# Hear my Voice: Understanding how community health workers in the Peruvian Amazon expanded their roles to mitigate the impact of the COVID-19 pandemic through Community-Based Participatory Action Research

**DOI:** 10.1101/2023.05.01.23289307

**Authors:** Tina Samsamshariat, Purnima Madhivanan, Alexandra Reyes, Eva M. Moya, Graciela Meza-Sánchez, Stefan Reinders, Magaly M. Blas

## Abstract

**Introduction:** The COVID-19 pandemic led to the collapse of the Peruvian health system, which disrupted healthcare access for indigenous communities in the Amazon. We aimed to understand how the COVID-19 pandemic transformed the responsibilities of community health workers (CHWs) from indigenous communities in the Peruvian Amazon so policymakers can support indigenous health efforts.

**Methods:** Fourteen CHWs from Loreto, Peru participated in a community-based Participatory Action Research (CBPR) project using Photovoice, a technique that encourages vulnerable groups to take photos and develop stories illustrating their lived experiences. Participants were recruited from *Mamás del Río*, a local university-based program, through purposive sampling. CHWs were trained in Photovoice and asked to photograph how the pandemic affected their lives and work. Participants met four times over five months to share photos and develop action items. Data were organized into key themes using a general inductive method. Final photos and action items were shared with policymakers during galleries in Iquitos and Lima.

**Results:** CHWs took a total of 36 photos with 33 accompanying texts highlighting their roles during the pandemic. Four core themes emerged: (1) the collapse of social infrastructure, (2) the use of medicinal plants versus pharmaceuticals, (3) the community adaptations and struggles, and (4) the importance of CHWs. CHWs expanded their responsibilities or leveraged their leadership across these themes to support COVID-19 patients, vaccination, and mandates without training or resources from the government. CHWs asked policymakers for formal integration into the health system, standardization of CHW training, and better management of community pharmacies.

**Conclusion:** CHWs, who work on a voluntary basis, took on additional roles during the pandemic with little to no training from the government. CHWs demonstrated how their roles could be better supported by the government to ameliorate future health catastrophes in the Peruvian Amazon.

**Short Summary:** Health care delivery in the Peruvian Amazon collapsed during the COVID-19 pandemic. Community health workers provided frontline care, education, and logistical support without formal training, resources, or compensation from the Ministry of Health. The government can better leverage, supervise, recognize, and integrate the role of community health workers to strengthen the health system.

## Introduction

Peru reported its first case of COVID-19 (SARS-CoV-2) in March of 2020 (1). The government issued a nationwide lockdown 10 days after the first case of COVID was reported (2). Despite the initial lockdown, systemic and social factors such as an underfunded and fragmented health system, in addition to water and electricity shortages, a diverse migrant population, and multigenerational housing, made containing the virus a challenge for the government (3,4). By 2021, Peru had proportionally the world’s highest COVID-19 death toll in the world at 5,551 COVID deaths per million (5).

Globally, indigenous populations have been disproportionately affected by the COVID-19 pandemic as measured by economic, cultural, or mortality indicators (6,7). In Latin America, mistrust of authorities, COVID-19 misinformation, lack of clean water, and unemployment created additional barriers for indigenous populations to combat the pandemic (8). Loreto, an Amazonian department in northeast Peru with the highest number of registered indigenous communities in the country, became one of the hardest hit regions during the pandemic (1). Some health centers chose to close early in the pandemic because of the lack of personnel, equipment, and training to manage the health crisis (9).

Despite the disruption to the health system and inconsistent protocols, two-thirds of community health workers registered with *Mamás del Río* (MDR), a program based at Universidad Peruana Cayetano Heredia (UPCH), continued to promote the health of their communities during the pandemic (9). Community health workers (CHWs) are legally recognized as “those people chosen or recognized by their community, who carry out voluntary actions for health promotion and disease prevention, in coordination with health personnel and other institutions (10).” CHWs were globally recognized as a crucial component to primary health care delivery by the 1978 Declaration of Alma Ata (11). The CHW system in Peru is currently fragmented with CHWs receiving training from the Ministry of Health (MINSA), religious congregations, and non-governmental organizations.

Given how CHWs from indigenous communities in the Peruvian Amazon continued to provide care despite the disruption of health services, we aimed to understand how the pandemic affected their responsibilities. Photovoice, a visual research methodology that empowers vulnerable groups to stimulate social change, was used to investigate how CHWs expanded their roles and mitigated the effects of the pandemic (12). Photovoice empowered CHWs to identify gaps in the health system through photography and to present their solutions to policymakers, who in turn could better support indigenous health in future disasters.

## Methods

### Study Design

Photovoice was used to understand how CHWs in the Peruvian Amazon adapted their lives and work to mitigate the health and social effects of the COVID-19 pandemic in their communities. Photovoice is a form of community-based, participatory action research (CBPR) that transcends qualitative research by generating empowerment and social change among participants through photography, group discussion, and reflection (13). The three main goals of photovoice are (1) to enable people to record and reflect upon their community’s strengths and concerns, (2) to promote critical dialogue about important issues through group discussion of photographs, and (3) to reach policymakers (14,15). Photovoice has been used with marginalized communities around the world, including immigrants and refugees experiencing social challenges such as living with HIV, mental illness, or homelessness (13). After participants identify community strengths and challenges through photos and discussion, they are given the opportunity to share their work with policy leaders. Thus, the researcher plays a passive role in the development of themes and solutions as the participant takes ownership of the project.

Photovoice was considered an appropriate methodology for the study because it allowed CHWs from vulnerable communities to share their experiences during the pandemic, to develop solutions to the challenges they face as CHWs, and to present their message to policymakers from the Health Sector who could address those challenges. Participants were divided into two groups and explored research questions posed both by researchers and themselves (Table 1). These questions included:

> **Groups A and B**. How did the COVID-19 pandemic affect my life and work as a CHW?
>
> **Group A.** During the pandemic, what was my work as a CHW to promote the health and wellbeing of my community?
>
> **Group B**. What is my work and responsibility as a CHW during the pandemic to promote the health of the community?

**Table 1.** Meeting Sessions and Objectives

| Meeting # | Objective | Research Question Explored | Question Development | # Of Photos Discussed |
| --- | --- | --- | --- | --- |
| 1 | Orientation to photovoice, general training in methodology, collecting informed consents, and ethics | None | None | None |
| 2 | Discussion of photos and group theme development | **Nauta (Group A) & Parinari (Group B):** How did the COVID-19 pandemic affect my life and work as a community health worker? | Question predefined by research team | Nauta: 14 |
|  |  |  |  | Parinari: 9 |
| 3 | Discussion of photos and group theme development | **Nauta:** During the pandemic, what was my work as a CHW to promote the health and well-being of my community? | Question developed by each group of participants | Nauta: 14 |
|  |  | **Parinari:** What is my work and responsibility as a CHW during the pandemic to promote the health of the community? |  | Parinari: 17 |
| 4 | Develop Call to Action and Finalize Photo Gallery | None | None | None |

### Study Setting

This study took place over a five-month period from February to June 2022. The study was conducted among two independent groups of CHWs in the districts of Nauta (Group A) and Parinari (Group B). These districts are two of the 53 districts in the department of Loreto, Peru (Figure 1, (16–20)). Loreto is one of the poorest regions in the country, with a poverty rate between 36.7% - 40.9% (21). More than one-third of its population lives in rural settlements connected by the river system with little to no land access (22).

**Figure 1.**
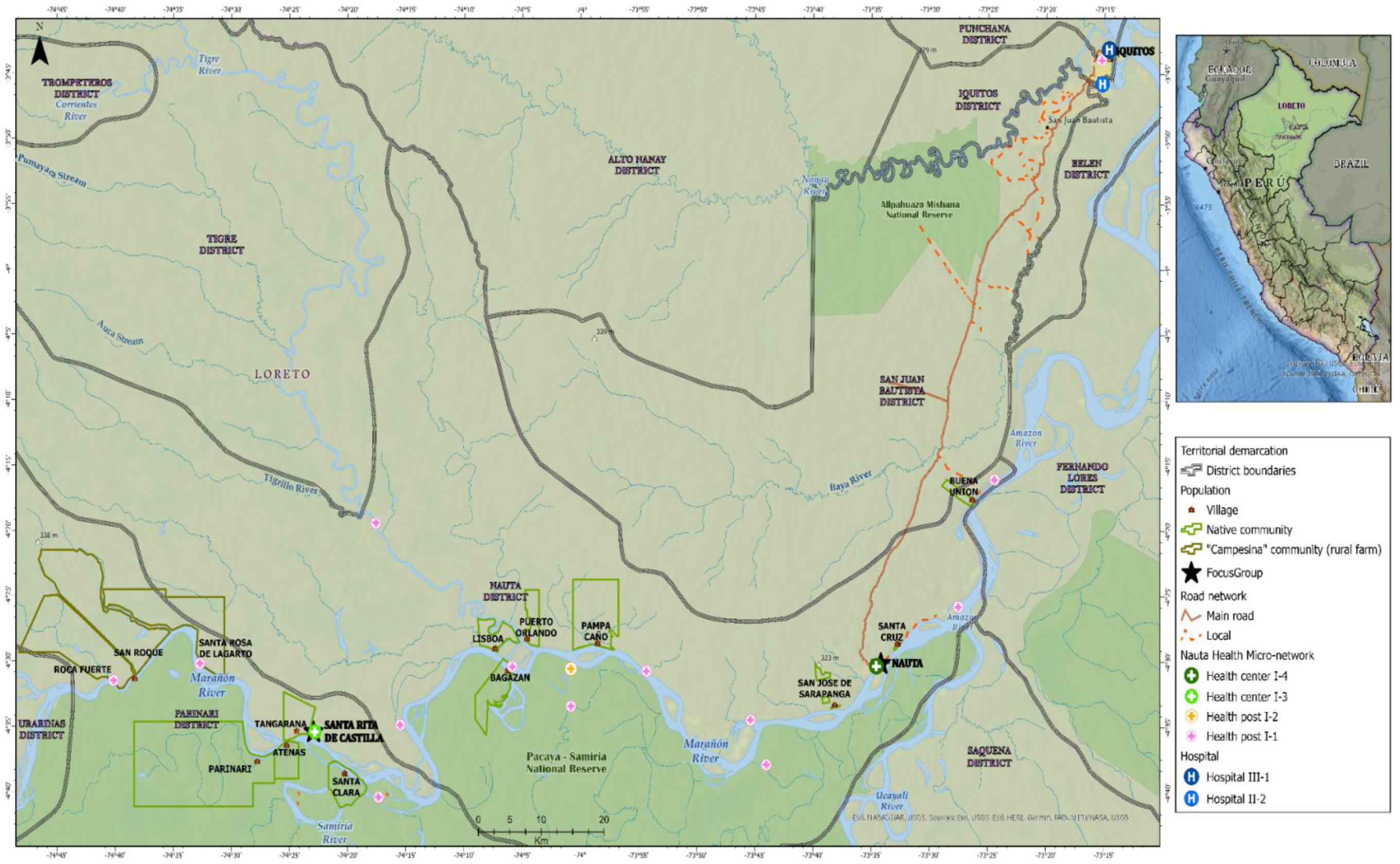
Map study location. Inclusion criteria included having served as a CHW through the COVID-19 pandemic, having the ability to provide informed consent, and having lived in the districts of Nauta or Parinari. All participants were required to attend the initial training session to remain in the project. All participants identified as belonging to the *Kukuma-Kukamiria* ethnic group. Their main activities include fishing and agriculture based on their close relationship with the rivers (29–31).

### Study Context

This study was conducted in partnership with *Mamás del Río* (MDR) with support from the Regional Health Agency (GERESA) of Loreto, previously known as DIRESA. MDR is a community-based, maternal, and neonatal health program through UPCH (9,23–27). MDR trains CHWs from rural communities in Loreto to conduct home visits and promote essential newborn care and healthcare seeking behavior among women from pregnancy to their postpartum period. Each district is made up of multiple communities with one CHW elected to represent each community. MDR held one training session on COVID-19 prevention and vaccination for their CHWs.

### Study Participants

A total of 20 CHWs were contacted to participate in the study. We aimed to recruit 10 participants from Nauta and 10 participants from Parinari based on sample sizes from previous photovoice studies (28). Participants were recruited from *MDR* through purposive sampling to reduce bias. Key sampling features included sex, length of time serving as a CHW, and time of travel to the

### Patient and Public Involvement

This study was designed and implemented in partnership with *Mamás del Río*, UPCH, GERESA-Loreto, GOREL-Loreto, the Ministry of Health (MINSA), the Pan American Health Organization, and indigenous communities. All partners were involved in protocol development and project delivery. Partners served as key stakeholders, recruited participants, and contributed to outcome measures. Community consultation and permission were acquired from community health workers, community authorities, and health centers. Participants played a key role in defining the research question and approving all research conclusions. Project findings were disseminated to communities via photo albums that contained all project content and acknowledged participants for their contributions.

### Procedures

The procedure was developed based on a typical nine step Photovoice approach as highlighted in Table 2 (14,15). The study was split between the districts of Nauta and Parinari because of the burden of travel, to keep discussion groups intimate, and to obtain a geographically diverse sample. Procedures were identical among both groups. The Nauta group met inside a church parish, and the Parinari group met inside a cultural building in Santa Rita de Castilla. The study sites lay 134 km apart (5-10 hours by boat, depending on river conditions) along the Marañón River. Figure 1 illustrates all participating communities and their relation to the primary study sites. The fieldwork team was composed of two primary researchers: TS (principal investigator) and AR (anthropologist), and a logistics coordinator. Researchers commuted between study groups once a week by boat.

**Table 2.**
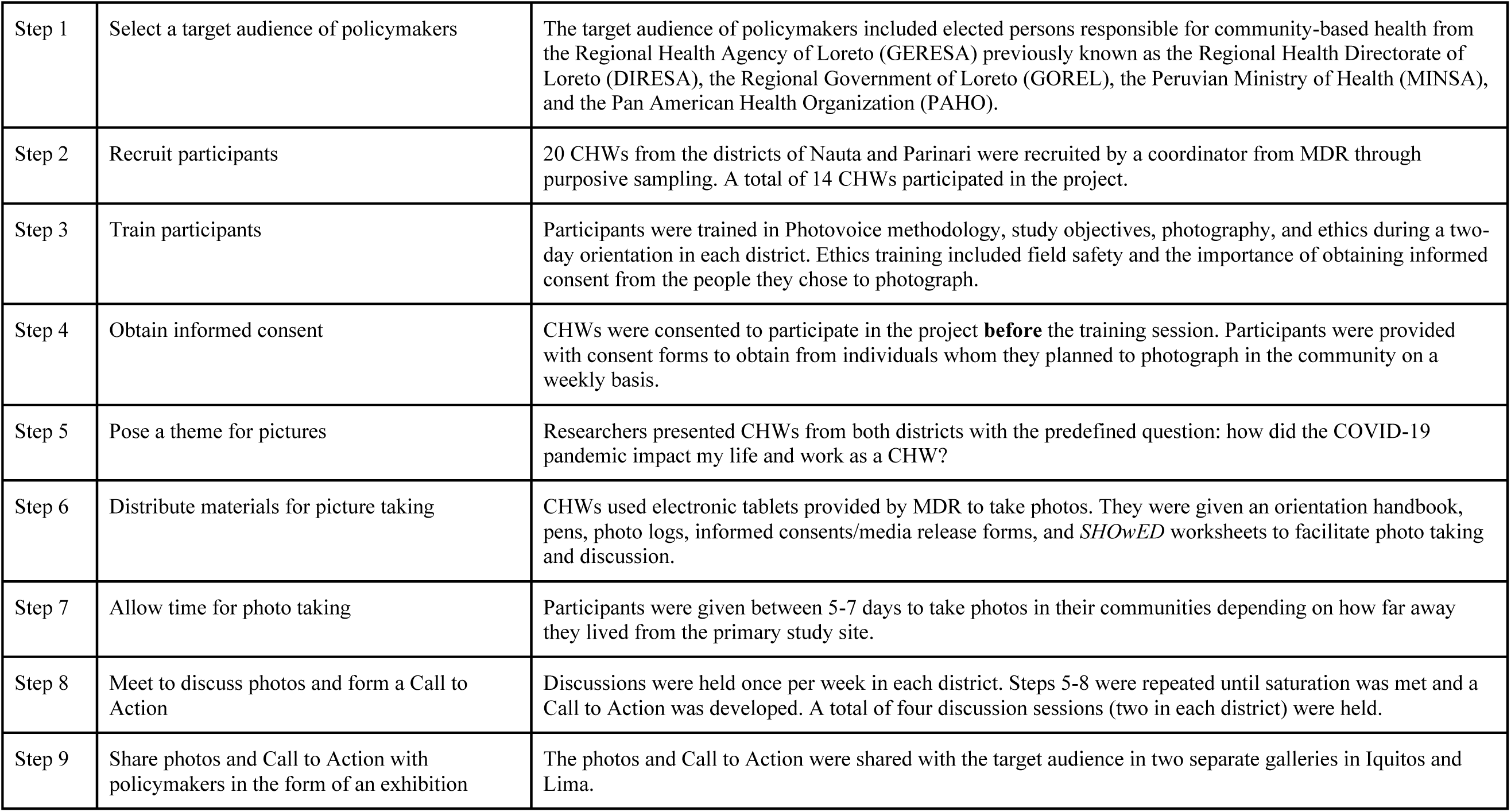
Steps outlining the typical approach to a photovoice project as applicable to this study

|  |  |  |
| --- | --- | --- |
| Step 1 | Select a target audience of policymakers | The target audience of policymakers included elected persons responsible for community-based health from the Regional Health Agency of Loreto (GERESA) previously known as the Regional Health Directorate of Loreto (DIRESA), the Regional Government of Loreto (GOREL), the Peruvian Ministry of Health (MINSA), and the Pan American Health Organization (PAHO). |
| Step 2 | Recruit participants | 20 CHWs from the districts of Nauta and Parinari were recruited by a coordinator from MDR through purposive sampling. A total of 14 CHWs participated in the project. |
| Step 3 | Train participants | Participants were trained in Photovoice methodology, study objectives, photography, and ethics during a two-day orientation in each district. Ethics training included field safety and the importance of obtaining informed consent from the people they chose to photograph. |
| Step 4 | Obtain informed consent | CHWs were consented to participate in the project **before** the training session. Participants were provided with consent forms to obtain from individuals whom they planned to photograph in the community on a weekly basis. |
| Step 5 | Pose a theme for pictures | Researchers presented CHWs from both districts with the predefined question: how did the COVID-19 pandemic impact my life and work as a CHW? |
| Step 6 | Distribute materials for picture taking | CHWs used electronic tablets provided by MDR to take photos. They were given an orientation handbook, pens, photo logs, informed consents/media release forms, and *SHOwED* worksheets to facilitate photo taking and discussion. |
| Step 7 | Allow time for photo taking | Participants were given between 5-7 days to take photos in their communities depending on how far away they lived from the primary study site. |
| Step 8 | Meet to discuss photos and form a Call to Action | Discussions were held once per week in each district. Steps 5-8 were repeated until saturation was met and a Call to Action was developed. A total of four discussion sessions (two in each district) were held. |
| Step 9 | Share photos and Call to Action with policymakers in the form of an exhibition | The photos and Call to Action were shared with the target audience in two separate galleries in Iquitos and Lima. |

The target audience of policymakers was identified at the start of the project so they could serve as project stakeholders. Meetings were held with stakeholders over the course of the project to ensure their participation in the final gallery.

During the two-day orientation, participants were trained in Photovoice methodology and field safety. Demographic data were collected in the form of a 14-question survey. Researchers presented both groups with the primary question: how did the COVID-19 pandemic impact your life and work as a CHW? This was a broad question so it could be contextualized by participants.

*SHOwED*, a mnemonic in which participants create a concise story explaining their photos, was used during discussion sessions(14). Questions include: What do you **See** here? What really is **Happening** here? How does this relate to **Our** lives? Why does this situation, concern, or strength **Exist**? What can we **Do** about it? Participants shared two to three photos during weekly discussions. WhatsApp, an end-to-end encrypted messaging application, was used to share and transmit photos between participants and researchers.

After the first session, researchers only posed framing questions to CHWs during discussions. This empowered participants to take ownership of the project as they developed subsequent research questions and relevant themes. This also allowed themes to emerge organically from group discussions. The groups voted on one photo from each participant to include in the final gallery during each session.

Overall, there were a total of four meetings in each district: one training session, two focus group discussions, and one Call to Action meeting. All sessions were led by TS and AR in Spanish. All discussions were recorded and transcribed verbatim into Spanish by university anthropology students. Saturation was met after the second discussion session, where participants from both groups voted that they did not need any more photo taking sessions.

All English translations were first done by AR and then edited independently by TS. Any conflicts were jointly discussed until resolution. Translations were made without overt corrections to grammar or sentence structure to maintain the authenticity of speech.

### Consent

Written consents and photographic releases were obtained from each participant. Participation was voluntary. All participants were compensated 15 Soles ($4.02 USD) for each meeting session they attended. Participants received reimbursement for all travel expenses, overnight lodging, and three meals per day. Participants were made aware of study goals and objectives. No pseudonyms were used during meeting sessions. Participants were required to obtain written consent of any human subjects they chose to photograph over the course of the study.

### Data Analysis and Thematic Development

Transcripts were uploaded into Atlas.ti, an online software, for data analysis. The data from each group were first analyzed separately and then combined to form conclusions. A general inductive approach was taken. This analytical framework allows the researcher to evaluate qualitative data through three main steps: condensing raw data into codes, linking codes to form categories, and finally, linking categories to describe the most important themes (32). Photo units, consisting of the photo title, the photo(s), and the *SHOwED* text, were analyzed in conjunction with transcripts. A total of 33 photo units were analyzed. The themes identified in the field by participants were broken down into codes with their respective photo units and then linked back into categories and final themes by researchers. Codes were maintained in a shared codebook, and final themes were collectively agreed upon by TS and AR after multiple rounds of reading and re-reading texts.

The final meeting, named the “Call to Action” meeting, took place in April 2022. Researchers combined themes from both groups into a single photo gallery because of overlap. The combined gallery and final themes were approved by all participants for publishing in the exhibition.

## Results

Fourteen out of 20 CHWs (n=14) participated in the study, with seven participants belonging to the Nauta group and seven belonging to the Parinari group. Six CHWs were unable to attend the initial training session, which disqualified them from participating in the remainder of the study. One participant contracted COVID-19 and was excluded from the study. *Data Supplement 1* includes participant demographics.

A total of 28 photos were shared among the Nauta group, and a total of 26 photos were shared among the Parinari group. Thirty-six photos with 33 accompanying texts were voted to be included in the final combined gallery. Four core themes emerged: (*1*) *the collapse of social infrastructure,* (*2*) *the use of medicinal plants versus pharmaceuticals,* (*3*) *community adaptation strategies and struggles, and* (*4*) *the importance of the community health worker.* Photos and texts included in the manuscript were chosen by the research team.

### The Collapse of Social Infrastructure

The collapse of social infrastructure during the pandemic hindered community development. Most prominent was the collapse of the transportation and healthcare systems.

The primary form of transportation in this region is personal canoes known as *peque peques.* Lockdowns mandated early in the pandemic prevented residents from leaving or entering the communities. Because communities depend on the river system to exchange food supplies, buy and sell economic goods, and travel to the nearest health facilities during emergencies, the interruption of transportation devastated communities. Participants explained how they traveled in secret during the night to bring back food or medicines from the nearest port town. Communities were forced to break their own lockdown regulations because the State failed to provide basic supplies. One CHW shared the importance of the boats in transporting patients to health centers:

> *“It is our everyday mobility that helps us in any emergency: it allows us to save the lives of ourselves or neighbors. **During the pandemic, we transported our patients to the nearest health center.** In an emergency you can mobilize quickly [with a boat], and when you cannot, you have to look for a neighbor or someone else [with a boat], because there is no ambulance in our community. **This boat is very useful for everything in our daily lives. Without it, we do nothing, we can die.** We wish that the community has a speedboat or small canoe dedicated for the transport of patients in emergencies and to carry out procedures”* (CHW8, Parinari).

While select communities have their own health post, others need to travel to the nearest health post to receive care. Although the health centers in Nauta and Santa Rita stayed open, it could take from 30 minutes to two hours to reach these health centers depending on river conditions. One participant explained how the closure of the health posts placed greater responsibility on the CHW to provide basic care in the communities (Image 1):

**Image 1.**
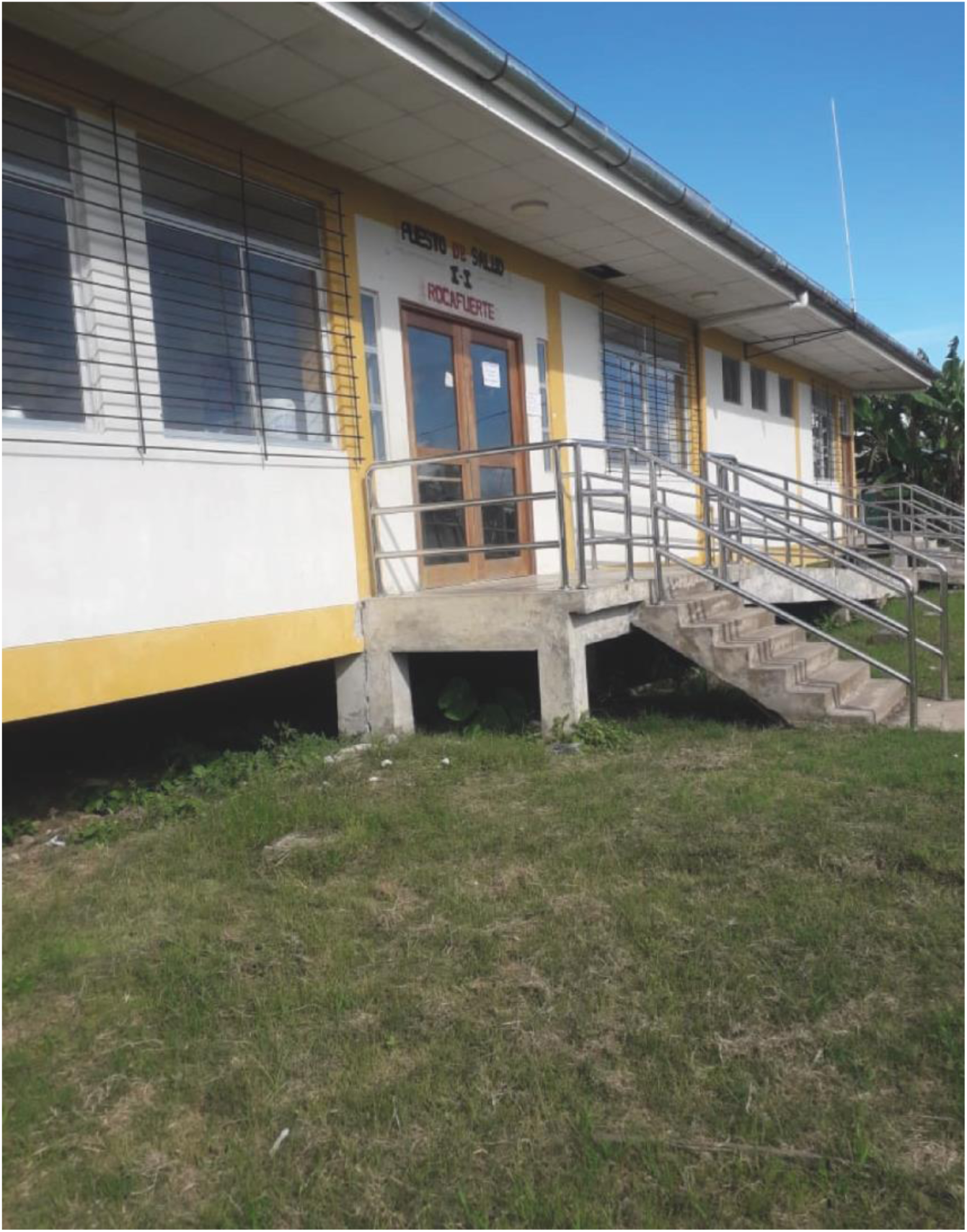
“The Health Post”

> *“The health post is closed because there was no medical care for COVID-19 in the communities. When the pandemic came, everything totally changed: they did not have medicines, nor did they have a full professional staff. We were afraid. Who remained when the post was understaffed or out of stock? That was the task of the community health worker. **I believe that our authorities, with the person-in-charge of the health post, can collaborate to make a bag of medicines that can be available to all the community members.** If we are an ally of the health establishment…they need to consider us, because where there are no posts in the communities, the community health worker intervenes”* (CHW11, Parinari).
>
> Despite the collapse of these social systems, participants generally felt optimistic that these systems would return to the same degree of stability as before the pandemic.

### The use of Medicinal Plants versus Pharmaceuticals

Due to the collapse of social infrastructure, communities were unable to access western medicines. A major emphasis was placed on the use of medicinal plants to cure patients of COVID-19. Grapefruit, *Sangre de Grado* (Dragon’s Blood), and *Cordoncillo* (spiked pepper) were used to treat COVID-19 patients. The knowledge of plant remedies was revered by all participants, and participants used the focus groups to exchange different plant recipes. Basic knowledge of medicinal plants was considered part of the community health workers’ responsibility.

Participants agreed that community members should each have their own plot to grow medicinal plants for emergencies. As one CHW explained,

> “*Since the pandemic is not over, we must continue to prepare those life-saving vegetables and eat them to protect ourselves. We should all have a well-organized garden with plants to take care of our health” (CHW2, Nauta)*.

In addition to their medicinal properties, these plants were also used as food and economic goods. One CHW explained the importance of the banana plant:

> *“We didn’t have money, everything we had was gone. **What did we do? Go to the plants, because the price of the pills has gone up and they couldn’t even sell us at the clinics**…without this plant, we would not have survived in times of COVID”* (CHW8, Parinari).

The collapse of supply chains prevented communities in the Amazon from accessing pharmaceutical drugs during the pandemic. Most communities have a “community medicine cabinet” or “communal pharmacy,” which is a small box containing essential medicines such as analgesics, nonsteroidal anti-inflammatory drugs, and blood pressure medications that are maintained by a community authority. These medicines are cyclically replenished by the regional health district or community donations. These cabinets remained empty during the pandemic because the regional health district could not send supplies due to the lack of transportation, and the high cost of medicine. One participant described how the lack of pharmaceuticals was a major barrier to health in the communities (Image 2):

**Image 2.**
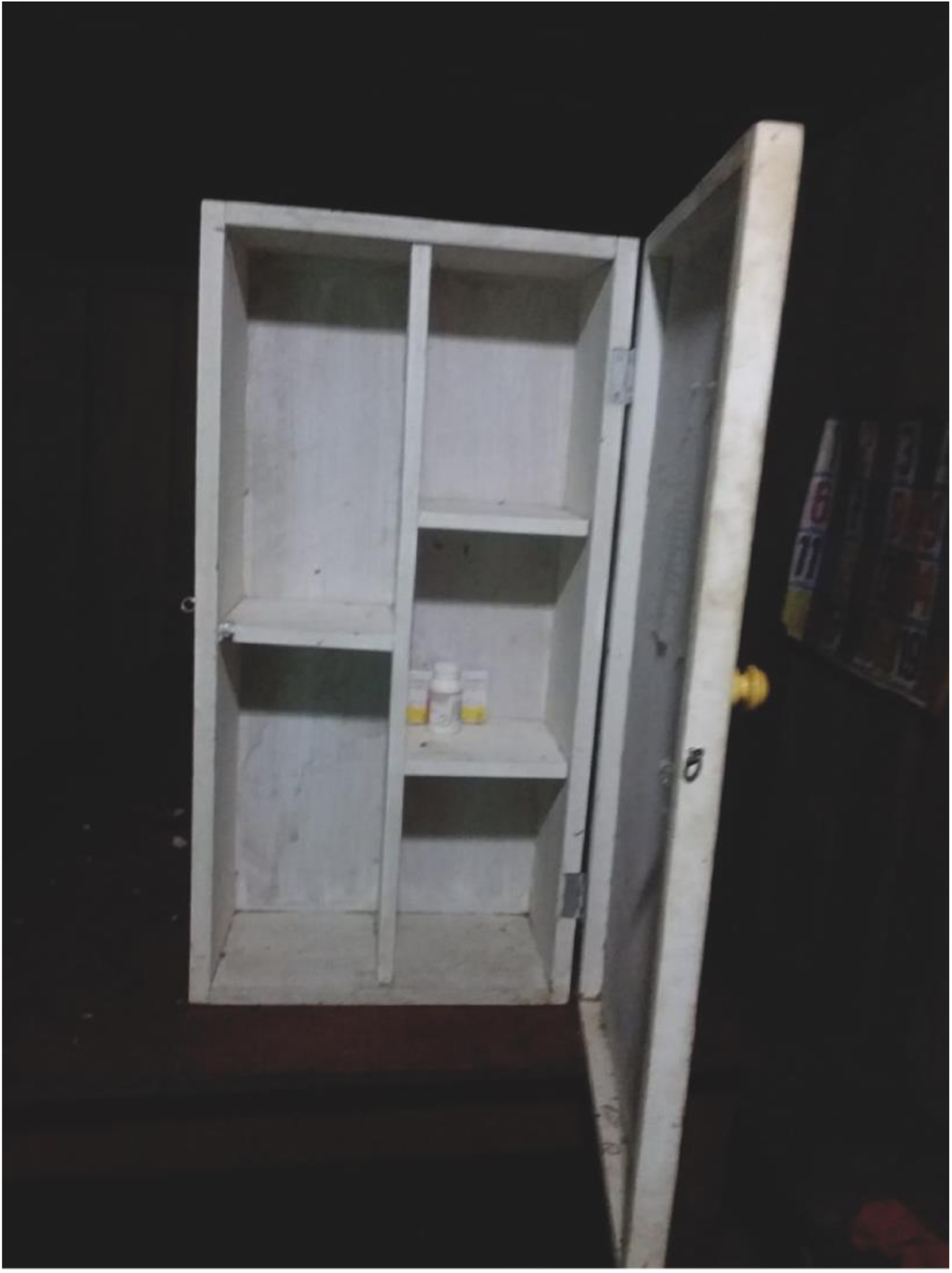
“The Community Medicine Cabinet”

> *“This little box is a community medicine cabinet. In the community, it is the responsibility of the health promoter, and it is mainly for the benefit of the community. As a CHW, I feel worried because it is empty. **It is like a soldier who goes to war without weapons**. You are left with great despair at not being able to provide support. The disease comes and we do not make it alive. Our role is not only to treat the patient, but also to support him so that he can reach the health post. Therefore, we must promote communal medicine cabinets with higher authorities. We want training on medicine administration so that we can be prepared for an emergency in our community”* (CHW6, Nauta).

### Community Adaptation Strategies and Struggles

The transition from a culture of togetherness to one of social distancing and isolation in accordance with COVID-19 protocols was difficult for many community members. As one participant from Parinari shared,

> *“We were not used to living locked up. We were used to living free”* (CHW10, Parinari).

Families needed to adapt to keeping their distance from sick relatives. One participant described the fear of leaving their family members alone at the health center.

> *“A pregnant woman in her bed is accompanied by her daughter. The pregnant woman is sick, she has discomfort throughout the body and a fever. The family is very concerned about COVID. **The State said that the sick patient with COVID has to go to places alone; but our custom says that the person cannot be alone. When you are accompanied, you feel safer, protected.** Care is the responsibility of the family. Several families had relatives arrive at the hospital, if they cured them, they would come out alive, otherwise, you would never see them. We have to organize ourselves in the community to manage a medical post with medications and trained professional nurses for emergency response and to prevent a pregnant woman or newborn from dying at any time”* (CHW9, Parinari).

The pandemic left communities with feelings of sadness, indifference, and vulnerability. One CHW described how communities were underprepared for these emotional struggles,

> *“Many of us have lost our family members: our father, our mother, our partner, children, grandfathers, grandmothers, neighbors. We have been unprepared and distrustful, thinking that nothing is going to happen. We have to help each other so that our families do not die from these diseases”* (CHW9, Parinari).

Despite the emotional struggles, participants generally remained hopeful they would overcome the pandemic by working together. Participants felt that COVID-19 protocols needed to continue to be enforced in the communities to prevent the further spread of the illness at the time of study.

### The Importance of the Community Health Worker

CHWs acknowledged their importance in providing basic health services during the pandemic. Not only did most CHWs remain committed to their established role during the pandemic, but some CHWs went beyond this role to take on new pandemic-related responsibilities, for which they had received little to no training from the government. These activities included caring for COVID-19 patients and leading the communities through COVID-19 vaccination. One participant shared his commitment to his patients:

> *“A community health worker is visiting a community member with COVID-19 in the middle of the pandemic. At first, I was afraid, but at that time, I would always check on [the patients] to see how they were, if they were getting better [or] if they were getting worse–that was my job. **My will is great. I don’t earn a penny; you don’t earn any incentive, but I work for my community.** There was a danger that some community members would die from COVID-19. Since I had protection, I would [make visits]. I was no longer afraid of death.”* (CHW2, Nauta).

CHWs highlighted how the State had failed to recognize their value to the communities. While this frustrated CHWs, it also served as a point of pride for their continued service. As one participant acknowledged,

> “*The concern is that there is a lack of support in this area from the government…they should worry about managing something [better] for the good of the people. We can help each other”* (CHW11, Parinari).

While many community members were eager to receive the vaccine, others were hesitant because of internet rumors. These rumors included that the vaccine was a government microchip, that the vaccine would make you infertile, that it would kill you, and that the vaccine was the “seal of the beast,” or the 666 seal from the Bible. One participant described how CHWs set an example for the community in lieu of these rumors:

> *“People were very desperate for vaccines. This concern existed because they told us that they did not have the remedies to cure us. **As community health workers, we must promote vaccination. We have an obligation so that other people can follow our example in the first, second and third doses.** We hear that “the vaccine is the seal of the beast; it has a chip.” We are key to showing why the vaccine is important, and that the first vaccine injected in the shoulder is the best vaccine. We can continue teaching and informing that getting vaccinated is best for everyone.” (CHW14, Parinari)*.

Not only did CHWs lead vaccine adoption, but they also helped organize the logistics of vaccine delivery with health posts and the *Apu*, or community chief.

### Call to Action and Photo Galleries

The Call to Action allowed participants to have a unified voice in asking for changes to improve healthcare delivery in the Amazon. The Call to Action asked policymakers for the formal integration of CHWs into the healthy system by:

1. Providing each CHW with an accreditation that certifies their role as part of the formal health system with remuneration.
2. Expanding and standardizing the role of CHWs by formally training them in first aid and other relevant health topics such as mental health and family planning.
3. Strengthening the community medicine cabinet in each community by sending essential medicines at least quarterly and training CHWs in the administration of pharmaceutical drugs.

A total of 36 photos with 33 associated texts were included in the gallery. A couple of participants decided to make photo collages, which accounts for the uneven number of photos and texts. Two photo exhibitions were held during the first week of June 2022, celebrating the “Day of the Community Health Worker,” which is recognized by the Ministry of Health (33). The first photo exhibition was held at the GOREL in Iquitos with local policymakers and CHWs. The second photo exhibition was held at MINSA in Lima with national government officials and broadcast online. A three-minute video showcasing the experiences of the CHWs during the pandemic and the Call to Action was presented to policymakers at both galleries.

## Discussion

Over the four-month period of photo taking, focus group discussion, and observation, participants shared how the COVID-19 pandemic affected their responsibilities as CHWs.

CHWs are legally defined as volunteers by the Ministry of Health, thus they work without any monetary incentive. The official responsibility of CHWs from *Mamás del Río* is limited to the surveillance and education of pregnant and recently delivered women in maternal and neonatal health. Despite the lack of infrastructure, compensation, or equipment to take on the challenges of the pandemic, CHWs exceeded their expected level of commitment by expanding their responsibilities in response to the shortcomings of the health system.

Four key themes emerged during this photovoice project: (1) the collapse of social infrastructure, (2) the use of medicinal plants versus pharmaceuticals, (3) community adaptation strategies and struggles, and (4) the importance of the community health worker. Participants expanded their scope of work directly or leveraged their leadership across these four themes during the pandemic.

Participants recommended standardized training in first aid, mental health, reproductive health, and medicine administration for all CHWs under the supervision of the health posts and MINSA. By training CHWs in general health topics and providing them with the tools to provide basic care in the communities, CHWs demonstrated that their roles could be expanded to buffer the collapse of health infrastructure. CHWs also highlighted how they used medicinal plants to care for COVID-19 patients when access to pharmaceutical medicines was severely restricted. CHWs are entrusted to care for sick individuals in the community regardless of why people are sick. Their knowledge and use of medicinal plants serve as a foundation for trust in their communities. This trust enshrines them as community leaders and important liaisons between the formal health system and indigenous health practices. This trust was critical in allowing CHWs to lead community adaptation efforts such as masking and vaccination as well as education on COVID-19 prevention and transmission. They were also responsible for coordinating vaccine delivery and transporting sick patients to the highest level of care available outside the communities. Lastly, they set an example for their communities in COVID-19 vaccination.

CHWs in other low- and middle-income countries (LMICs) have been facing similar structural challenges to healthcare delivery during the pandemic (34–38). The role of CHWs in LMICs, like Peru, needs to be refined to account for the pandemic setting. In countries with well-established CHW programs, such as Nigeria and Vietnam, the role of CHWs has changed to support community engagement, awareness, and contact tracing in the pandemic setting (39). When health systems protect CHWs, they can leverage their roles to interrupt virus transmission, maintain health care worker capacity, and shield the most vulnerable in society from the downstream socioeconomic effects of pandemics (40). CHWs can also provide psychosocial support to their communities to increase mental health services (41).

The Peruvian government does not currently recognize CHWs as part of the formal health system. This creates a structural barrier for CHWs in delivering coordinated and continuous care. Our study shows that CHWs in rural settings have the potential and desire to serve as frontline providers with the proper recognition, training, supervision, and tools. They also serve as a key cultural liaison between indigenous communities and the formal health system, which fosters trust between the State and community members (42). The role of CHWs as a bridge between the health system and vulnerable groups has been recognized and leveraged in Brazil, a South American country with an established CHW network (37).

Several study limitations must be addressed in the interpretation of these results. First, the small sample size (n=14) and the recruitment of all participants from a single university-based organization reduces generalizability to the experiences of other CHWs in Peru. Furthermore, researchers were non-independent from the organization, therefore results could be influenced by researcher bias. Generalizability is also limited as all participants identified as *Kukuma-Kukamiria*, which does not consider the experiences of other ethnic groups in this region. All participants lived within a reasonable distance to Iquitos by water or by land. Communities further away from the major cities have more limited resources and face additional logistical challenges that were not accounted for in this study.

The strength of using Photovoice in this setting is that it allowed participants to share their experiences directly with policymakers. All participants endorsed some level of increased empowerment at the end of the project, and many attributed their likening of Photovoice to the opportunity to share their voices. It was also a feasible modality in a rural setting with limited access to the internet or advanced technology. Further research is needed to explore how the role of the community health care worker can be standardized through training and remuneration in Peru. After the conclusion of this study, CHWs from MDR were empowered to create the first association of indigenous CHWs in Loreto called AACOSIL (Asociación de agentes comunitarios de salud intercultural de Loreto) with the objective to advocate for the recognition and inclusion of CHWs in the Peruvian health system.

## Conclusion

To our knowledge, this is the first study that explores the effects of the COVID-19 pandemic on the life and work of community health workers in the Peruvian Amazon through photovoice. Our findings add to the growing body of literature on the importance of training community health workers to address health gaps and health emergencies in LMICs. This study highlights the structural and logistical challenges faced by indigenous communities in combating the COVID-19 pandemic, in addition to the challenges faced by CHWs in mitigating the health crisis. CHWs in Loreto provided educational and logistical support during the pandemic as well as frontline care when health centers shut down without training, tools, or preparation. CHWs emerged as community leaders during the pandemic, and this leadership has the potential to be leveraged and supported by the Peruvian government to strengthen the health system.

## Data Availability Statement

Data are available upon request from the study PI (Tina Samamshariat)

## Ethics Statement

### Patient Consent for Publication

Consent obtained directly from patient(s).

### Ethics Approval

The study received approval from the institutional review boards of the UPCH (#207032) and University of Arizona (STUDY00000257).

## Data Availability

Data are available upon request from the study PI (Tina Samamshariat)

## Acknowledgements

We thank the dedicated community health workers in the Amazon that have continued to work through the pandemic. We thank the Pan American Health Organization for their support of the project. We acknowledge Camila Gonzales and Angela Alva for providing logistical support in the field. Diana Ayaucán Requena led graphic design. Jefferson Isla Ríos led video production. Karla Vergara Rodríguez authored geographic map. María Nieves and José Manuel translated all photo captions to *Kukama*. Naomi Bishop provided manuscript supervision.

## Author Reflexivity Statement

See Data Supplement 2.

## Supplementary Materials

### Supplementary Data

Data Supplement 1

Data Supplement 2

### Footnotes

**Twitter:** @Tsamamshariat, @Pmadhivanan01, @Gmezas, @MagalyBlasB

## Contributors

TS was responsible for study design, planning and facilitation of the intervention, data collection, analysis, interpretation, and write-up. PM was responsible for research mentorship, facilitation, and execution of study. AR was responsible for project planning, facilitation of intervention, anthropological consideration, data collection, analysis, interpretation, and translation. EM was responsible for project design and methodology. GMS was responsible for field coordination and community access. SR was responsible for project proposal, design, and write-up. MMB was responsible for mentorship, community access, project planning, intervention, and gallery design. TS, PM, and MB coordinated efforts for ethics approval through the University. All authors reviewed the final manuscript for publication.

## Patient and Public Involvement

Patients and/or the public were involved in the design, or conduct, or reporting, or dissemination plans of this research. Refer to the Methods section for further details.

