## Supplementary material for "Hear my Voice: Understanding how community health workers in the Peruvian Amazon expanded their roles to mitigate the impact of the COVID-19 pandemic through Community-Based Participatory Action Research": Data Supplement 1

Participant Demographic Characteristics

| **Participant characteristics** | Total (%)  (n = 14) |
| --- | --- |
| **District** | |
| Nauta | 7 (50%) |
| Parinari | 7 (50%) |
| **Sex** | |
| Female | 5 (36%) |
| Male | 9 (64%) |
| **Age (in years)** |  |
| 20-29 | 1 (7%) |
| 30-39 | 0 (0%) |
| 40-49 | 7 (50%) |
| 50-59 | 6 (43%) |
| **Ethnicity** |  |
| Kukama kukamiria | 14 (100%) |
| Other | 0 (0%) |
| **Education level** |  |
| None | 0 (0%) |
| Primary incomplete | 1 (7%) |
| Primary completed | 5 (36%) |
| Secondary incomplete | 5 (36%) |
| Secondary completed | 2 (14%) |
| Technical incomplete | 0 (0%) |
| Technical completed | 1 (7%) |
| Bachelor's incomplete | 0 (0%) |
| Bachelor's completed | 0 (0%) |
| **Time as a CHW** |  |
| Less than 1 year | 0 (0%) |
| Between 1 and 2 years | 0 (0%) |
| More than 2 years | 14 (100%) |
| **History of COVID-19 infection*** | |
| Yes | 11 (79%) |
| No | 3 (21%) |
| **Confirmation of COVID-19 infection by Test**** | |
| Yes | 5 (33%) |
| No | 9 (67%) |
| Prefer not to answer | 0 (0) |
| **History of COVID-19 Vaccine***** | |
| Vaccinated | 12 (93%) |
| Not vaccinated | 1 (7%) |
| **COVID-19 perceived as a threat** | |
| Yes | 13 (93%) |
| No | 0 (0) |
| Don’t know | 1 (7%) |

* Suspected infection by COVID-19 including those not confirmed by molecular or antigen test

** Antigen or molecular test

***At least two doses
