## Supplementary material for "Hear my Voice: Understanding how community health workers in the Peruvian Amazon expanded their roles to mitigate the impact of the COVID-19 pandemic through Community-Based Participatory Action Research": Data Supplement 2

**Author Reflexivity Statement**

1. **How does this study address local research and policy priorities?**

This original research was designed to specifically address gaps in healthcare for indigenous community members in the Peruvian Amazon. The work utilizes Photovoice, a visual research methodology, where participants defined the research question. Thus, the research team facilitated discussion and led analysis, but participants defined research objective and priorities. Furthermore, participants identified and presented policy concerns directly to local and national policymakers as a part of project methodology.

2. **How were local researchers involved in study design?**

The knowledge and involvement of local researchers has been crucial to the development of this project. MB serves as the last author of the paper, she is a physician-researcher based in Peru, who has devoted her career to indigenous health in the Peruvian Amazon. GM serves as a physician-policy maker based in Loreto, Peru. She provided policy support and access to communities. Both MB and GM have extensive research experience. AR is a trained anthropologist in Peru and guided all anthropological considerations of the project. PM, EM, TS, and SR have diverse cultural backgrounds; however, they represent high income country researchers on the work. They acknowledge their privileged positions in research and aim to promote equitable research for international colleagues.

**3. How has funding been used to support the local research team?**

This project received $10,000 USD from the US National Institutes of Health Global Health Equity Scholars Program and an additional $5,000 USD from the Pan American Health Organization. Funding has been used with local research team to purchase qualitative analytical software, graphic design software, and pay fair wages to research assistants. Funding was used to pay for all travel expenses of the local research team. Funding has also been allotted to give local research team the opportunity to travel and present the work to continue career growth.

**4. How are research staff who conducted data collection acknowledged?**

All research staff who conducted data collection are acknowledged as authors. Specific roles of authors are outlined in the acknowledgments.

**5. Do all members of the research partnership have access to study data?**

All team members have access to study data.

**6. How was data used to develop analytical skills within the partnership?**

Junior level researchers were required to complete the literature review and data analysis for the project. Additionally, all researchers were invited to attend a 2-day Photovoice workshop to learn more about the methodology and analyze this data set. Qualitative data analysis through synthesis and data coding were key analytical skills prioritized by the research team. Lastly, research team developed their language skills as transcripts and concepts were translated from Spanish to English.

**7. How have research partners collaborated in interpreting study data?**

Multiple virtual meetings were held with all research partners and local stakeholders during data interpretation. Key themes were identified through multiple rounds of meetings. All partners agreed on data interpretation in the context of respecting and promoting indigenous health.

**8. How were research partners supported to develop writing skills?**

Early career researchers (TS and AR) were supported by senior researchers through multiple revisions of the manuscript and all related writing through the project.

**9. How will research products be shared to address local needs?**

We wish to publish this work as open access. Additionally, all participants have received photo albums that contain the final photos, themes, text, and policy statements arising from the data analysis of this project. Participants have their own copies of their photos and all project deliverables to take back to their communities. The project was shared with policymakers at two galleries in Peru. Policymakers received copies of project deliverables to help their decision making in addressing local needs.

**10. How is the leadership, contribution and ownership of this work by LMIC researchers recognised within the authorship?**

MB (Peru) serves as the last author of the paper. She was the project mentor and primarily responsible for the leadership of the project. AR (Peru) and GM (Peru) serve as a middle authors and have been recognized in the acknowledgements for their contributions to the writing of this work and the overall project. Overall, this work has 7 authors. Three authors are from Peru and four authors are from high income countries. Although there are more authors from high income counties, local researchers and partners have been crucial to this work and recognized for their contributions to this project.

11. **How have early career researchers across the partnership been included within the authorship team?**

Early career researchers (TS and AR) have been heavily included in the development, analysis, fieldwork, and writing of this work. TS is a pre-doctoral MD student and served as the first author. AR is a young career anthropologist who served as a research assistant. She was heavily involved in fieldwork, data analysis, translation, and writing. Young career researchers attended methodology workshops and had the opportunity to attend all meetings with local and national policy makers.

**12. How has gender balance been addressed within the authorship?**

Six authors are female (TS, MB, PM, AR, EM, and GM) and one author is male (SR).

**13. How has the project contributed to training of LMIC researchers?**

Three authors (MB, GM, and AR) are LMIC researchers. MB and GM are senior level researchers. MB is the founder of Mamás del Río – the primary local partner. GM currently holds a director position for the Regional Health Government of Loreto. AR is a junior researcher. She was employed by project funding for the duration of the project. She took all anthropological leadership of the project and presented the work at the Ministry of Health and at national conferences.

**14. How has the project contributed to improvements in local infrastructure?**

After the project, participants were empowered to create the first federation of organized community health workers in the Peruvian Amazon. Their purpose is to continue advocating for appropriate integration, training, and support of community health workers in the Amazon by the Peruvian government. MB continues to work at the policy level through Mamás del Río to integrate community health into the formal health system. The objective of this project was to identify gaps in the community health worker system of Peru to address those gaps and improve health infrastructure.

**15. What safeguarding procedures were used to protect local study participants and researchers?**

During fieldwork, participants provided written consent to participate in the project. Additionally, all persons that were subjects of photos had to provide written consent for their photos to be taken. COVID-19 safety measures including social distancing, hand sanitization, and masking were taken during fieldwork. Participants who had photos included in the manuscript were also consented via the BMJ consent forms. All names and other data have been de-identified.
